# Mobility restrictions as a pandemic response

**DOI:** 10.1101/2022.02.11.22270865

**Authors:** Hakan Lane, Mehmet Şahin, Philipp Otto

**Author notes:** Corresponding Author: Hakan Lane.

## Abstract

The COVID19 pandemic has caused a large number of infections and fatalities, causing administrations at various levels to use different policy measures to reduce viral spread by limiting public mobility. This paper analyzes the complex association between the stringency of restrictions, public mobility, and reproduction rate (R-value) on a national level for Germany. The goals were to analyze; a) the correlation between government restrictions and public mobility and b) the association between public mobilities and virus reproduction. In addition to correlations, a Gaussian Process Regression Technique is used to fit the interaction between mobility and R-value. The main findings are that: (i) Government restrictions has a high association with reduced public mobilities, especially for non-food stores and public transport, (ii) Out of six measured public mobilities, retail, recreation, and transit station activities have the most significant impact on COVID19 reproduction rates. (iii) A mobility reduction of 30% is required to have a critical negative impact on case number dynamics, preventing further spread.

## 1. Introduction

The COVID-19 virus first appeared in China in December 2019 and spread rapidly around the world. The World Health Organization (WHO) declared it a Public Health Emergency of International Concern in January 2020 and as a pandemic on March 11 [1]. As of the end of May 2021, it has cost 3.5 million lives worldwide [2-3].

The airborne nature of the virus meant that it was imperative to ensure that infected persons spread it to a few others as possible. The pivotal average number of other individuals that each carrier of the virus infects at a given point in time is defined as the reproduction value (R-value) [4-5]. The goal of lowering the R-value made it essential to reduce close contact between individuals, and most policy responses targeted social distancing and public mobility reduction. This was pursued through various means by the national governments. Various countries or specific regions around the world have placed a wide variety of mobility restrictions to protect their populations from being infected. The efficiency of these measures can be quantified as to their isolated effect on changing the R-value.

The most successful restrictions appear to have been based on the premise of reducing a human to human contact. Through prohibitions, public closures, and recommendations, citizens were forced or encouraged to spend less time near others and more time in a private sphere, while keeping the maximum size of private gatherings at minimum levels. Note that all jurisdiction policies could be based on national, regional, or local level, and R-values are compiled and reported by government authorities in each nation. An international review of government interventions found that a combination of measures implemented at the right points in time was essential for curbing reproduction numbers [6].

General compliance with these policies could be estimated by observing the amount of traffic at public spaces of various kinds. For example, the current mobility could be measured through automated data collection implemented on a variety of platforms [7]. This allows for an estimate of the number of people passing at certain locations and is usually presented as daily aggregates.

Due to the variety in responses and reporting, the present study concentrates on the effects of policy implementation in a single country and use Germany as a case study for the dependencies between public policy, aggregate mobility, and virus contamination. According to the WHO, there were 3.7 million cases and 90,000 deaths in Germany through May 26, 2021. There were several national “lockdown” periods in Germany with a combination of measures from the list enforced by work-at-home orders, mandatory closures of public and private establishments, and travel restrictions [8]. The government response is related to loss of mobility and the spread of infections.

The nonlinear nature and influence of external factors in the relationship between mobility and virus transmission make modeling their interaction a complex task. While several conventional statistical approaches have been attempted [9-11], previous research shows indications that Gaussian Process Regression (GPR) models form a solid basis for models of virus spread [12,13]. In direct comparisons between the methods on COVID-19 data, GPR has been able to fit the data better than other machine learning methods like Support Vector Machines (SVM) or Decision Trees (DT) [14]. Due to its capacity to capture spatio-temporal variations combined with external factors, it can be considered the state of the art for modeling the geographical spread of diseases. This has been found also for other diseases such as malaria [15]. GPR seems to work well with small to moderate-sized datasets as in the COVID-19 pandemic (daily observations over slightly more than a year). The model was applied with a particular focus on threshold levels of mobility for the urgent goal of keeping R-values below one to dampen the spread of the virus. In this context, the contributions of the present study can be summarized as follows. First, the connection between stringency and mobility is quantified, identifying what sectors of public life are affected most by public restrictions. Second, the association between various mobilities and spread of the virus is established through correlation analyses, determining what mobilities has the most influence on the reproduction number. Finally, machine-learning analysis of mobility, vaccinations and temperature as predictors of virus spread leads to an estimate of 30 % as a threshold to contain further spread of pandemic infections.

## 2. Method

### 2.1. Data Sources

Google Mobility reports measures public mobility as aggregated averages of the number of observed individuals passing multiple monitored locations. Utilizing data from the Google Maps system and other platforms, it measures the amount of mobility in the categories in six categories presented in Table 1. The numbers are reported as the percentage change from a baseline level.

**Table 1.** Public mobility data categories.

| Category | Inclusion |
| --- | --- |
| Retail and Recreation | restaurants, cafes, shopping centers, theme parks, museums, libraries, and movie theaters |
| Grocery and Pharmacy | grocery markets, food warehouses, farmers markets, specialty food shops, drug stores, and pharmacies |
| Transit stations | subway, bus, and train stations |
| Parks | local parks, national parks, public beaches, marinas, dog parks, plazas, and public gardens |
| Workplaces | places of work |
| Residential | homes |

The government-level of restrictions is quantified in a project known as the Oxford COVID-19 Government Response operated by the Blavatnik School of Government at the University of Oxford. It incorporates a wide variety of different restrictions related to social distancing in the wake of the pandemic and serves as a benchmark on how much each administration enforced lockdown. The Stringency Index is a weighted average of several categories [16], including school closures, workplace closures, cancellation of public events, restrictions on public gatherings, closures of public transport, stay-at-home requirements, public information campaigns, restrictions on internal movements, and international travel controls.

Temperature data were collected from the German Weather Service (DWD). The daily average at the Berlin-Tegel station was used as an aggregate for each day in the studied period.

R-values were obtained from the Robert Koch Institute derived from a now-casting model which has been used to forecast virus propagation on national and local levels [17]. Through time series analysis of the number of new cases per day, an instantaneous reproduction number can be derived retrospectively for each day [18].

### 2.2. Mobility Correlations

Spearman correlations were used to identify associations between:

a. Oxford Stringency Index and the various mobilities: a positive association means that higher stringency led to higher mobility, whereas negative correlations indicate that restrictions served to reduce public movement.
b. Mobilities and reproduction values: positive associations would lead to the conclusion that increased mobilities caused wider spread.

### 2.3. Gaussian Process Regression

An exponential–logarithmic model has been identified as an adequate fit for the association between community mobilities and reproduction rates [19-20], i.e. the logarithm of the R-value is dependent on aggregate mobility. It has also been identified that both temperature and level of vaccinations have an impact on reproduction [21-22].

Gaussian Process Regression (GPR) is a nonparametric supervised machine learning method usually applied to multivariate classification and regression problems [23]. GPR is used for describing the original distribution for flexible classification and regression models, where regression or class probability functions are not only simple parametric forms. One of the main advantages of the Gaussian process is the diversity of covariance functions that leads to the formation of functions with distinct types or degrees of continuous structures and enables choosing the proper selection.

Based on these previous findings it was possible to fit a GPR model for the relationship between mobility, temperature, and vaccinations with the following setup:

Kernel function: Exponential

Kernel scale: 14.396

Signal standard deviation: 0.230

Sigma: 0.230

Training data: 80 % of observations, randomly chosen, Test data: Remaining 20 %

## 3. Results

### 3.1. Government stringency

The Oxford Stringency Index calculates stringency as a weighted index of government response related to nine factors: school closures; workplace closures; cancellation of public events; restrictions on public gatherings; closures of public transport; stay-at-home requirements; public information campaigns; restrictions on internal movements; and international travel controls.in four dimensions. Estimated levels of stringency of German policy are on display in Figure 1, on a scale from 0 to 100 %.

**Figure 1.**
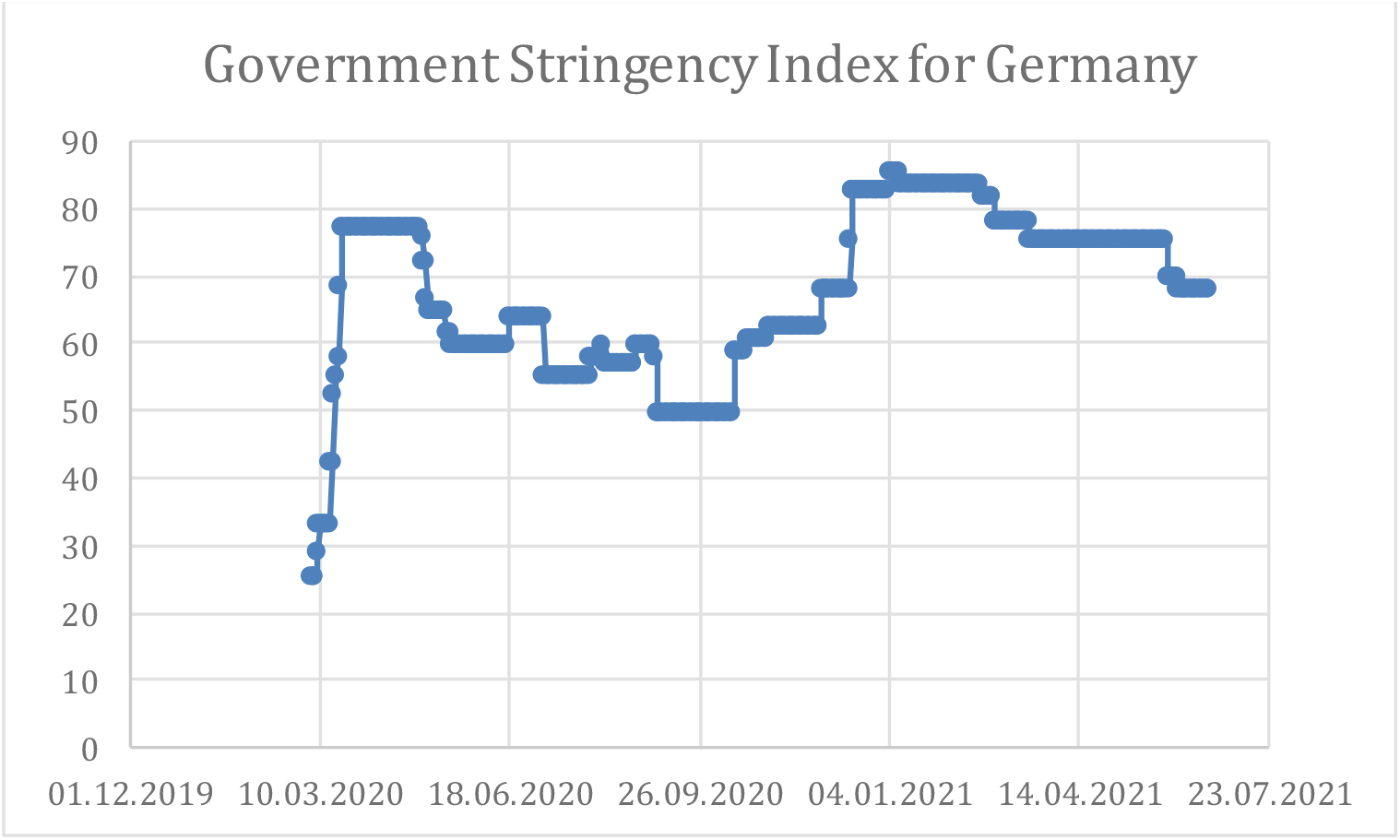
Government Stringency.

### 3.2. Public mobilities

Public mobilities measured in percentage change from the baseline in the six categories (Table 1) are in Figure 2.

**Figure 2.**
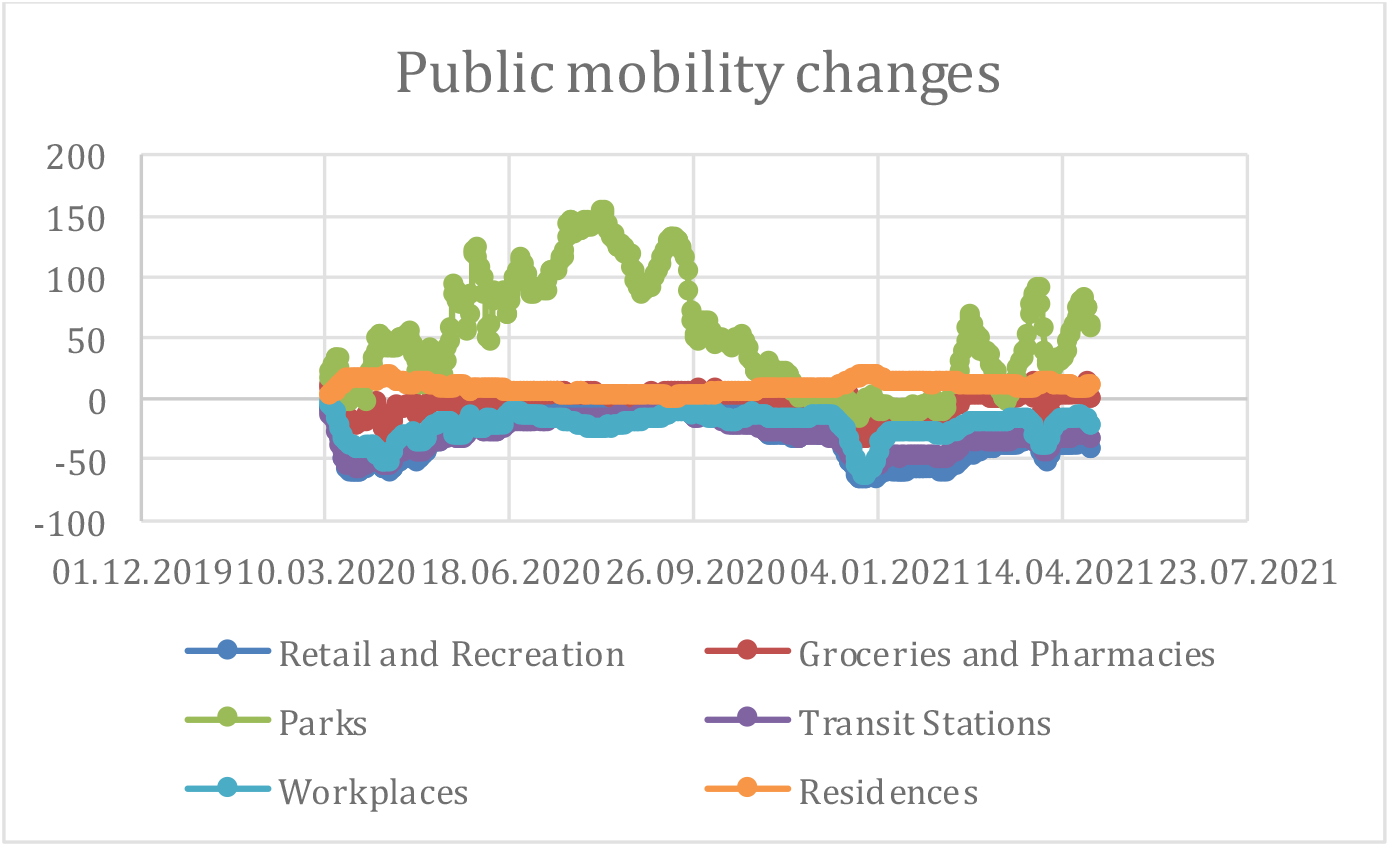
Public mobilities.

Activity at workplaces, retail and recreation, and public transit stations was well below ordinary levels through the period, while parks rose during the Summer and residential mobilities was higher due to more work from home arrangements.

### 3.3. Correlations Analysis

To compare the efficiency of the implemented policy measures and evaluate what sectors had the most impact on virus propagation, an attempt to quantify the association between government stringency, R-value, and each of the six mobilities. Spearman correlations between daily values of overall stringency and public mobilities are presented in Figure 3 and Figure 4, respectively.

**Figure 3.**
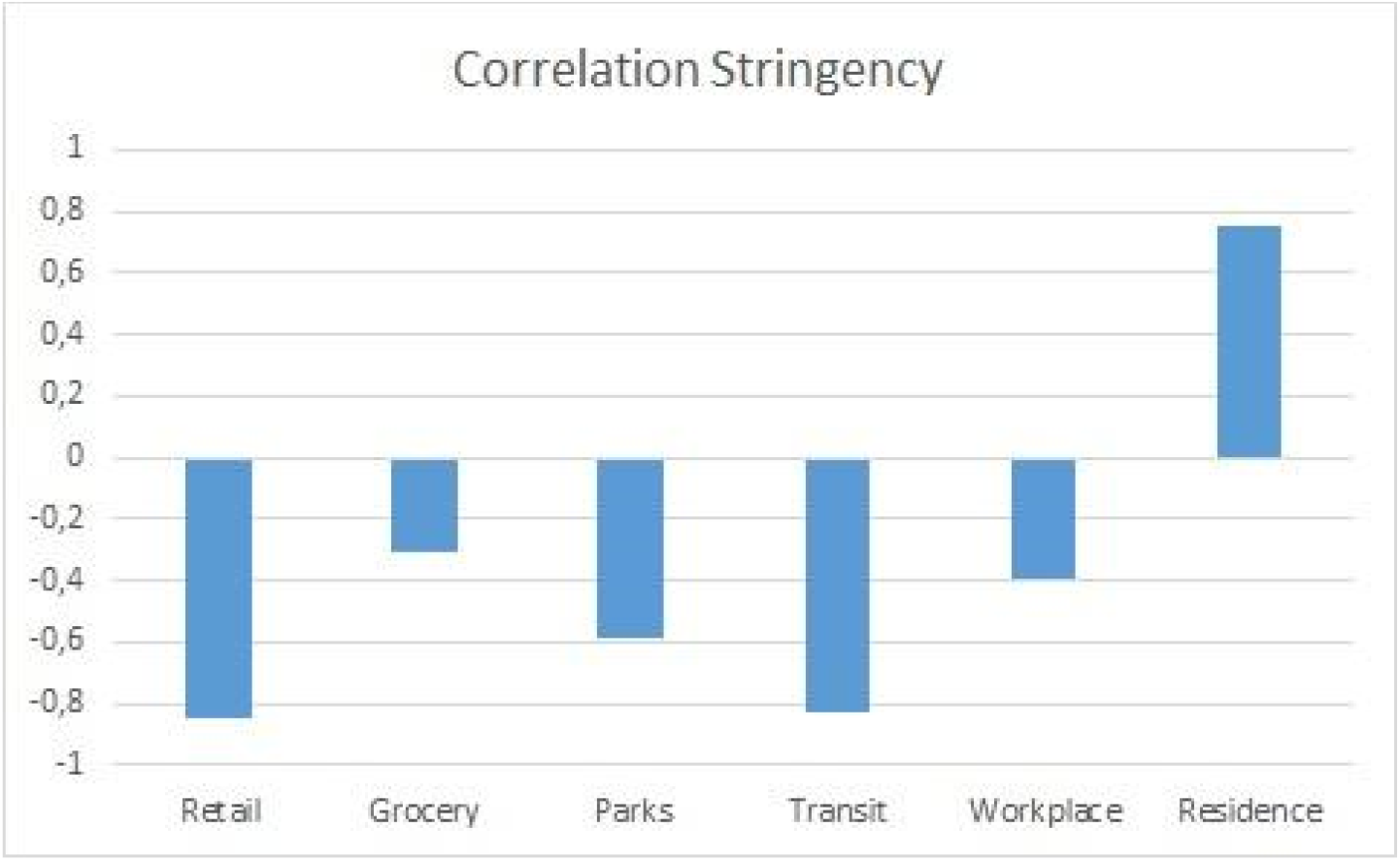
Correlations Stringency – Mobility.

**Figure 4.**
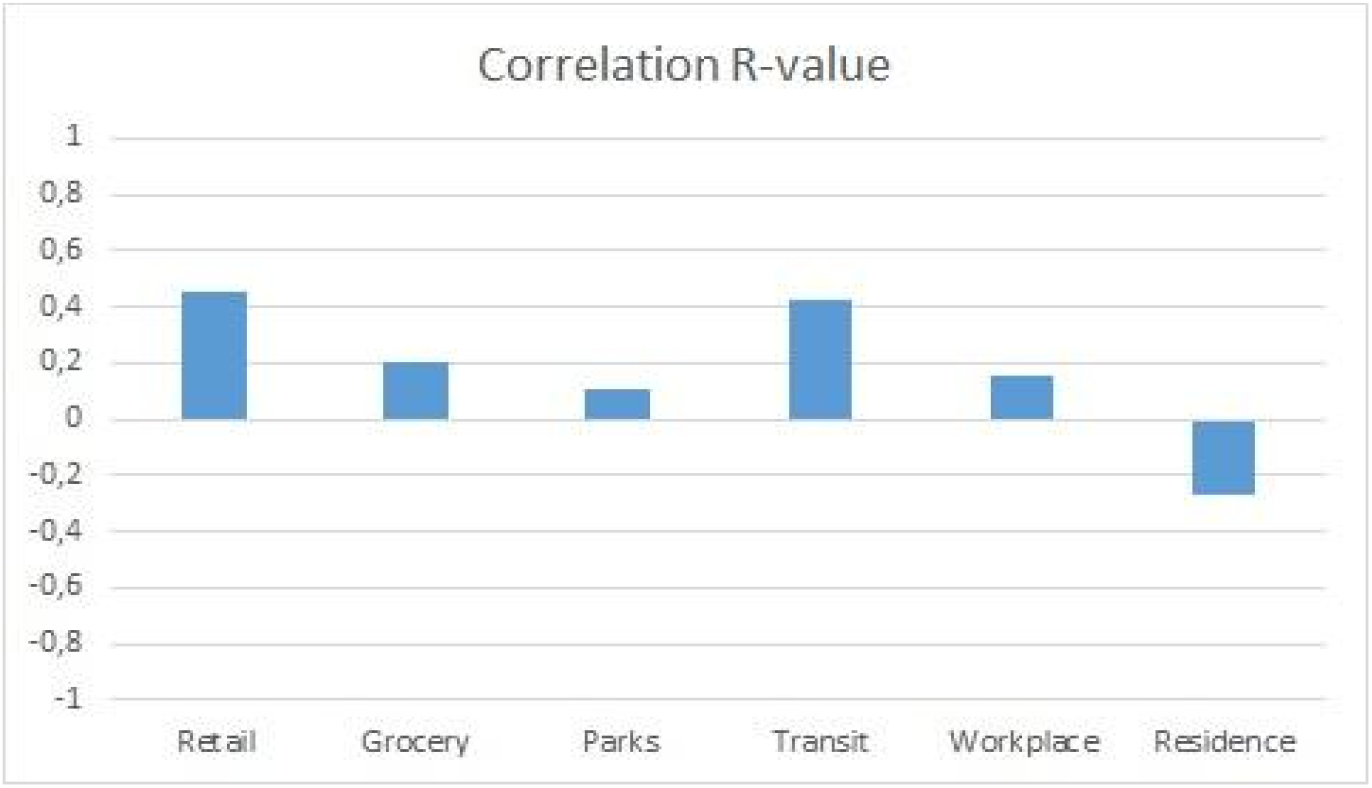
Correlations sector mobilities and reproduction.

A higher amount of stringency had the strongest negative impact primarily retail and transit sectors, while more restrictions also were associated with more time spent at home. The clearest associations were between mobilities in the retail and transit sectors, respectively, where increased levels of mobility led to higher R-values. More time spent in the residence slowed virus reproduction, while especially parks and workplaces had a rather small impact.

### 3.5. GPR Model Fit

An evaluation of the model fits of GPR compared to linear regression on the test set is presented in Table 2 and the prediction accuracy plot is given in Figure 5.

**Table 2.** GPR Model fit results

| RMSE | MAE | MSE | R2 |
| --- | --- | --- | --- |
| 0.14 | 0.093 | 0.018 | 0.83 |

RMSE=Root Mean Square Error; MAE=Mean Absolute Error; MSE = Mean Square Error; R2=Coefficient of Determination

**Figure 5.**
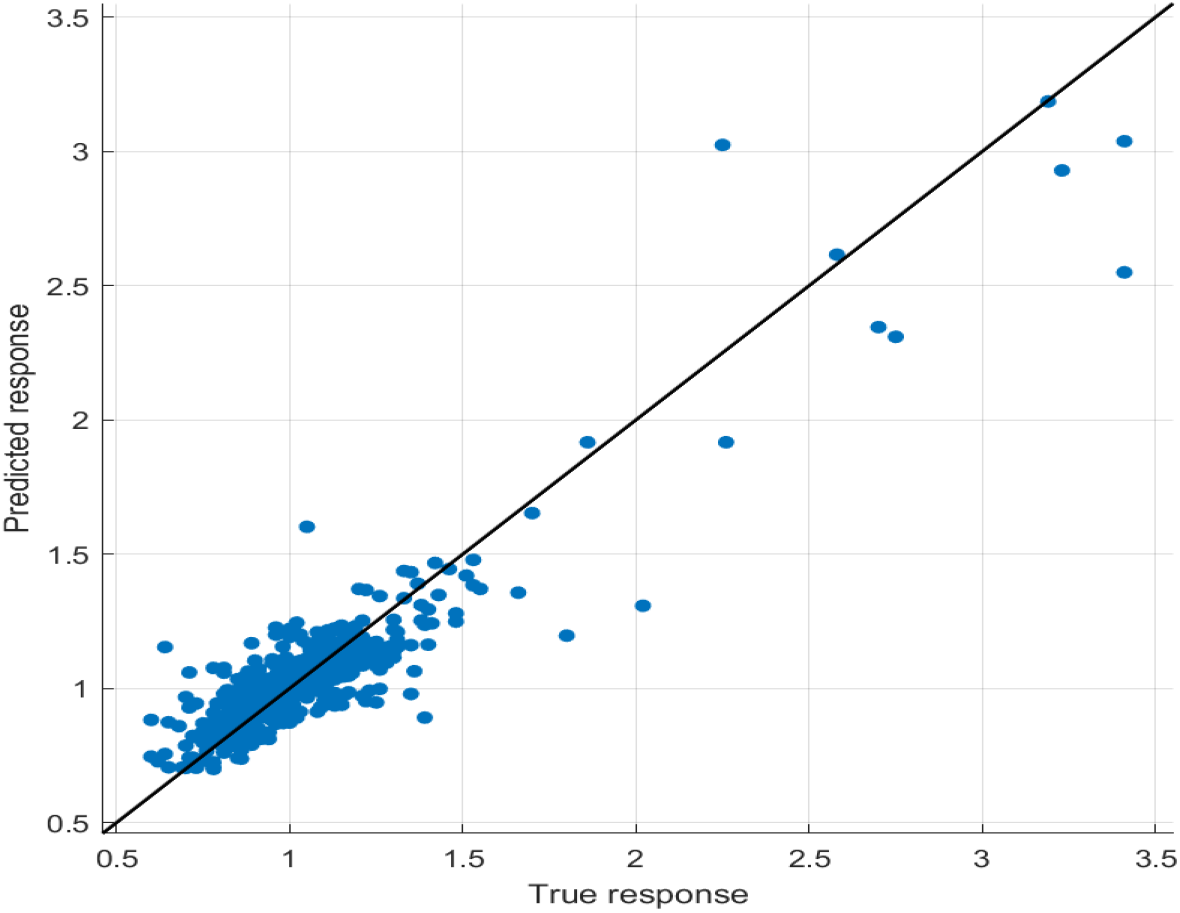
Model fit of GPR model.

The GPR with an exponential kernel captures the modeled relationship more accurately than the conventional method as it leads to a closer fit and lower prediction errors, along with a coefficient of determination above 80 %.

### 3.4 Mobility Thresholds

The reproduction rate was associated primarily with variations in mobilities for (i) retail and recreation (ii) transit stations. Based on this, it was possible to estimate the probability for an R-value under one for each range of mobility reductions. The results are displayed in Figure 6 and Fig. 7, respectively.

**Figure 6.**
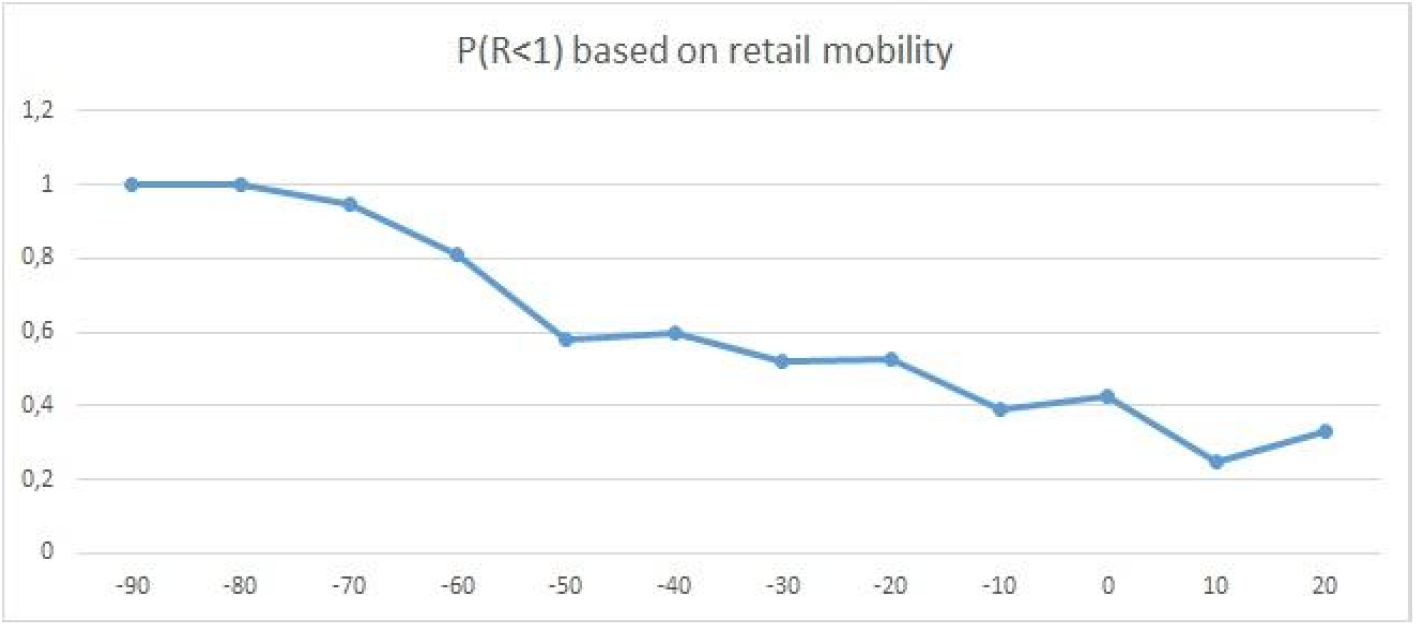
Estimated probability of R below spread levels based on retail mobility.

**Figure 7.**
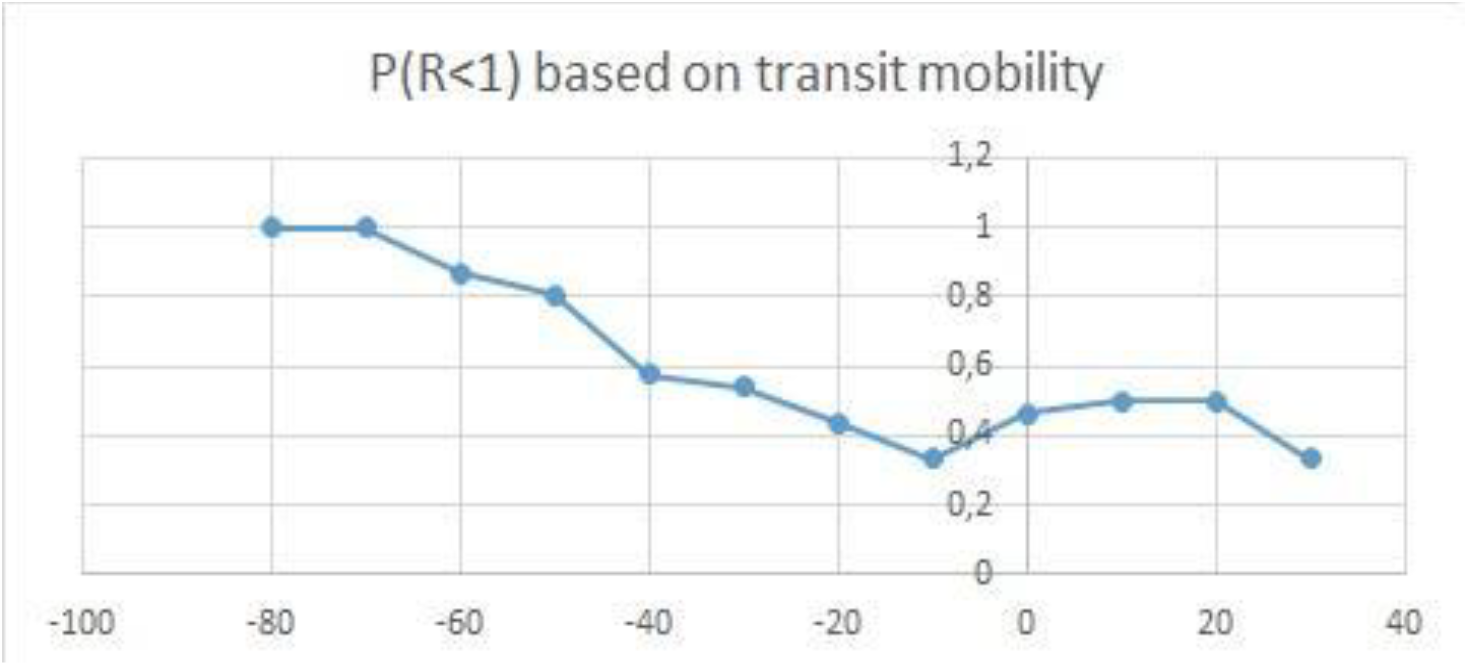
Estimated probability of R below spread levels based on transit station mobility.

Both graphs indicate a 30% reduction of mobility as a threshold for a higher than 50 % probability of R being at levels below 1 preventing further spread.

## 4. Discussion

The highest correlation between mobility and reproduction was identified for retail and transport, as these sectors seem to be the most pivotal for containing the virus. These would be perceived as non-essential and tend to gather considerable amounts of human beings in small inside spaces, especially during peak hours. These results still need to be confirmed beyond the German example, where other behavioral patterns might be more prevalent. The goal of restricting community spread by containing R-values below one with at least 50% chance required a reduction of 30% in retail or transit mobilities, similar to previous results [36].

It was expected that higher mobilities would lead to more reproduction and that vaccination would drive the number in the opposite direction, matching common theories on virus propagation. The negative association with temperature was also in line with anticipations, as the virus is known to spread at higher rates in lower temperatures. As indicated by the high rise in parks mobility in the Summer months, people are also spending a higher share of their time outdoors during warmer periods, making them less prone to infect other members of the public.

## 5. Conclusions

Understanding the relationship between stringency, mobility, and infection rates have been crucial for coping with the worldwide outbreak of COVID-19, and data from Germany for the period from March 2020 to May 2021 served as an example. Higher correlation between activity at retail/recreation and transit locations and virus propagation points to public transport and cultural meeting points as areas of high importance for mitigating the pandemic. It appears that the German official strategy to contain these by closing a large number of cultural and recreational establishments and reducing utilization of public transport through stay-at-home orders was well-founded, while activities at workplaces, parks, and grocery stores had less impact.The targeted level of 30% should serve as a benchmark threshold for necessary mobility reductions in future similar outbreaks.

## Data Availability

All data produced in the present study are available upon reasonable request to the authors

https://ourworldindata.org/covid-mobility-trends

## Data Availability

Government Stringency Index values are publicly available through Our World in Data at https://ourworldindata.org/covid-stringency-index.

Public Mobility values can be found through the Imperial Yougov behavioral tracker data repository at https://github.com/YouGov-Data/covid-19-tracker.

Nowcasting values of the reproduction number R were obtained through the Robert Koch Institut https://www.rki.de/DE/Content/InfAZ/N/Neuartiges_Coronavirus/Projekte_RKI/Nowcasting.html.

